# Incidence of frailty-related fracture among Medicaid beneficiaries living with HIV and cancer: A cohort study

**DOI:** 10.1101/2025.11.24.25340893

**Authors:** Xueer Zhang, Jacqueline E. Rudolph, Yiyi Zhou, Xiaoqiang Xu, Keri L. Calkins, Eryka L. Wentz, Bryan Lau, Corinne E. Joshu

## Abstract

**Background:** People living with HIV (PLWH) are at increased risk for frailty-related fracture. Limited evidence suggests recent diagnosis of non-AIDS defining cancer (NADC) is a risk factor for fracture among PLWH. We evaluated frailty-related fracture by HIV and cancer status to inform the burden of fracture among PLWH.

**Methods:** We included 14,554,711 beneficiaries without HIV and 159,188 beneficiaries with HIV who were 30-64 years old enrolled in Medicaid between 2001-2015 in 14 states. HIV, NADC, and fracture diagnoses were identified from inpatient and other non-prescription claims. We calculated age-specific fracture incidence per 100 person-years and 95% confidence intervals for beneficiaries with no NADC or HIV, NADC only, HIV only, or both. We estimated the cumulative incidence of frailty-related fracture with death as a competing event for each group.

**Results:** Fracture incidence increased with age in all groups. Compared to beneficiaries without NADC or HIV, all groups had significantly higher age-specific incidence of fracture. Beneficiaries with HIV and NADC had higher incidence of frailty-related fracture than those with HIV only at all ages (Incidence per 100 person-years for ages 30-44 Both: 1.24 95%CI:0.96,1.59; HIV: 0.74 95%CI:0.71,0.77; for ages 60-64 Both: 2.11 95%CI:1.64,2.67; HIV: 1.47 95%CI:1.37,1.58). Conversely, cumulative incidence was lowest for both HIV and NADC, likely due to in part to the very high incidence of death.

**Conclusion:** Beneficiaries with HIV and NADC had higher age-specific frailty-related fracture and death than beneficiaries with HIV alone. Future work should investigate fracture risk among PLWH with cancer to inform interventions for fracture prevention.

## INTRODUCTION

Frailty-related fractures are a significant comorbidity among people living with HIV (PLWH) in the modern era of antiretroviral therapy.^1–5^ Evidence suggests that PLWH have a higher risk of frailty-related fractures than the general population and that these may occur at earlier ages.^6^ Risk factors for frailty and related fractures, including pelvic, vertebral, hip, and wrist fractures,^1–5^ include older age, smoking, osteoporosis, or HCV co-infection.^7^ Higher fracture incidence is of particular concern given incident fracture has been associated with a nearly 50% increased risk of all-cause mortality among PLWH.^3^

Cancer is also a significant comorbidity among PLWH.^8–10^ In the general population, cancer survivors have an increased risk of frailty-related fractures, particularly those treated with chemotherapy and/or hormonal therapies, which adversely affects bone mineral density.^11–16^ Little is known about fracture risk in the complex scenario of HIV infection and cancer, but a prior study reported that a recent non-AIDS defining cancer diagnosis was associated with a significantly increased fracture risk among PLWH.^17^

To expand this work and better inform care for PLWH, we evaluated the burden of frailty-related fractures among PLWH with cancer compared to those with HIV only, cancer only, or neither condition. We included more than 14 million Medicaid beneficiaries 30-64 years, enrolled in 14 states, and between 2001 and 2015. Medicaid, a joint federal and state program, provides insurance coverage to individuals that meet certain low-income and disability-related eligibility criteria, including approximately 40% of US PLWH.^18,19^ We included beneficiaries <50, the age at which frailty-related fracture increases among PLWH,^6,20^ to assess whether a diagnosis of cancer contributes to fractures at earlier ages.

## Methods

We evaluated claims data from Medicaid beneficiaries enrolled from 2001-2015 in 14 US states: Alabama, California, Colorado, Florida, Georgia, Illinois, Maryland, Massachusetts, New York, North Carolina, Ohio, Pennsylvania, Texas, and Washington. Eligible beneficiaries were aged 30-64 years with full benefits and ≥7 months of continuous coverage without dual enrollment in Medicare or private insurance to avoid missed diagnoses of dual enrollees.^21^ We excluded beneficiaries <30 years due to low frailty-related fracture prevalence. We included beneficiaries’ first eligibility period of ≥7 months to allow for a 6-month run-in period to ascertain medical histories and reduce capture of prevalent outcomes and ≥ 1 month of subsequent follow-up. Of the 15,555,927 eligible for analysis, we excluded beneficiaries with any cancer (n=479,642) or fracture (n=362,386) ICD-9 diagnosis code (Table 1S) during the run-in period. The final analytic sample included 14,713,899 beneficiaries. The secondary data used in this analysis was accessed between 5/5/23 and 5/14/24. The Johns Hopkins Bloomberg School of Public Health Institutional Review Board approved and waived the requirement for informed consent for this analysis of a Limited Data Set, which contains protected health information including dates of birth, death, and healthcare services.

We classified beneficiaries as having HIV or cancer if they had one inpatient claim or two outpatient claims within 2 years with an HIV-related (during the run-in period or follow-up period) or cancer-related (during follow-up period) ICD-9 diagnosis code, respectively (Table S1).^22^ We used the first claim date as the date of diagnosis. We included non-AIDS defining cancers (NADC), which were previously associated with increased fracture risk among PLWH.^17^ We also assessed the cumulative incidence of frailty-related fracture for AIDS-defining (ADC), lung, colon, breast (female only), and prostate (male only) cancers separately. All beneficiaries were classified into four time-updated, exposure categories: no HIV or cancer, HIV only, cancer only, or both HIV and cancer.

We identified the first occurrence of frailty-related fractures (hip, radial, vertebral, or pelvic)^14,15,23–27^ during the follow-up period. We classified beneficiaries as having a fracture if they had one inpatient or outpatient claim with a fracture-related ICD-9 diagnosis code (Table S1) during the follow-up period. ^28^ We used the first claim date as the date of fracture.

Baseline was defined as the later of two dates: the day a beneficiary reached age 30 or the first day following the 6-month run-in period. For analyses stratified by age (30-44, 45-49, 50-54, 55-59, or 60-64) or calendar year period (2001-2005, 2006-2010, or 2011-2015), follow-up time began at baseline or on the first day of the age/calendar year period, whichever happened later. From baseline, beneficiaries were followed until: (1) fracture diagnosis, (2) death, (3) lapse in Medicaid coverage to avoid potential missed diagnoses during the lapse, (4), reached age 65, or (5) end of ICD-9 coding (September 30, 2015), whichever happened first. For analyses stratified by cancer type, beneficiaries were additionally censored at diagnosis of any other cancer.

We identified beneficiaries’ age, sex (male or female), race/ethnicity (non-Hispanic white or non-Hispanic Black), state and calendar year of enrollment at baseline. We calculated the medians and proportions of beneficiary characteristics by HIV and cancer status. We calculated the proportion of the first frailty-related fracture by body location (hip, pelvic, vertebral, or wrist) by HIV and cancer status. We calculated age-category-specific crude incidence rates of fracture and death as the number of events per 100 person years at risk by HIV and cancer status. We used the mid-P test to estimate their 95% confidence intervals.^29^ We used the Aalen-Johansen estimator to estimate cumulative incidence curves for fracture and death by HIV and cancer status using age as the time scale.^30^ Death was treated as a competing event. Analyses were carried out overall and stratified by sex. All analyses were conducted using R version 4.3.3.

## Results

Our analysis included 14,554,711 Medicaid beneficiaries, with 159,188 PLWH at baseline. The median age was comparable among beneficiaries with (43.9 yrs) and without HIV (43.5 yrs). PLWH had a lower proportion of female (PLWH: 34.7%, No HIV: 57.1%) beneficiaries and a higher proportion of Black (PLWH: 47.8%, No HIV: 20.6%) beneficiaries as compared to those without HIV (Table S2).

The incidence rate of frailty-related fractures increased with age among all categories of beneficiaries overall and by sex (Figure 1; Tables S3,S4). For all age groups, beneficiaries with HIV, NADC, or both all had significantly higher incidence rates of frailty-related fracture than beneficiaries with none of these conditions. These differences were the largest in the youngest age group, 30-44 years, and attenuated slightly with age. As compared to beneficiaries without either condition, age-specific incidence rates were 2-3 times higher for beneficiaries with both HIV and NADC, 1.4-1.8 times higher for beneficiaries with HIV only, and 2-2.4 times higher for beneficiaries with NADC only.

**Figure 1.**
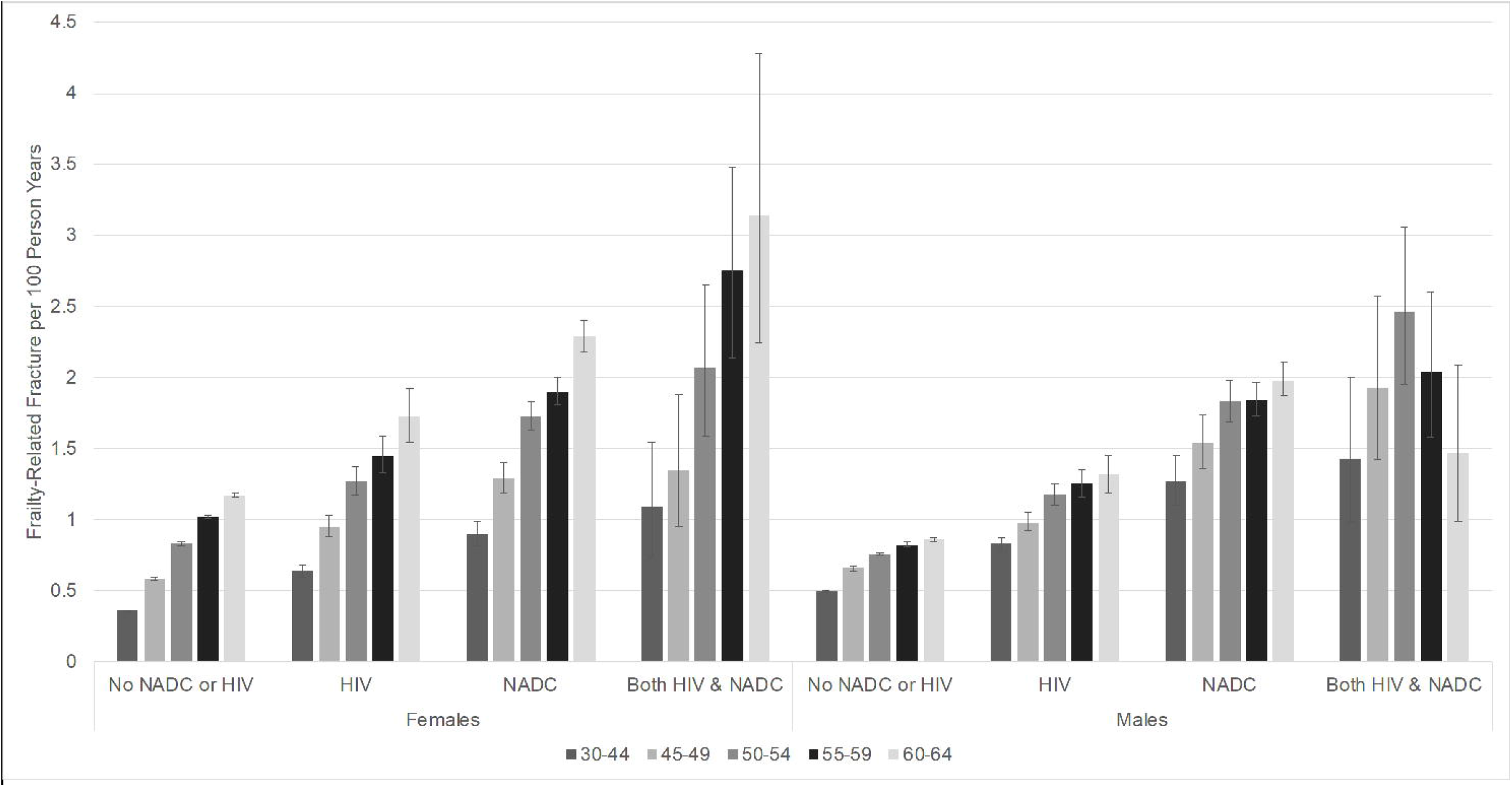
Crude incidence rates of frailty-related fracture per 100 person years by sex, age, and HIV and non-AIDS defining cancer (NADC) status among Medicaid Beneficiaries, 2001-2015. Error bars: 95% confidence intervals.

Wrist and hip fractures were the most common first fractures. Wrist fractures decreased with age and hip fractures increased with age except among female beneficiaries with HIV and NADC (Table S5). Vertebrae factures were generally consistent across age, but more common among male beneficiaries as compared to their female counterparts. Pelvic fractures were the least common fracture for all groups.

When comparing males and females, among beneficiaries without either condition, males had higher incidence of frailty-related fractures than their female counterparts at younger ages, but females had a higher incidence beginning at age 50 (Figure 1, Table S4). For all other groups, males and females generally had similar incidence of frailty-related fractures except for age 60-64 years, in which female beneficiaries had higher incidence than their male counterparts.

Age-specific incidence of frailty-related fracture was generally highest among beneficiaries with both HIV and NADC, followed by beneficiaries with NADC only, beneficiaries with HIV only, and then beneficiaries with neither condition (Figure 1; Tables S3,S4). Beneficiaries with both HIV and NADC had significantly higher age-specific incidence of frailty-related fracture than beneficiaries with HIV only at all ages (Table S3). This pattern was similar for females and males, though not statistically significant for female beneficiaries 50-54 and male beneficiaries 55-59 and 60-64 (Figure 1, Table S4). Beneficiaries with both HIV and NADC had non-significantly higher age-specific incidence of frailty-related fracture than beneficiaries with NADC only overall and for females and males, except for males 60-64 (Figure 1; Tables S3,S4).

When assessing the cumulative incidence of frailty-related fracture and death, these patterns were largely similar except for beneficiaries with both HIV and NADC, who had the lowest cumulative incidence of frailty-related fracture, particularly at older ages (Figure 2A,C,E). This finding was likely influenced by the substantially higher cumulative incidence of death among beneficiaries with both HIV and NADC (Figure 2B,D,F).

**Figure 2.**
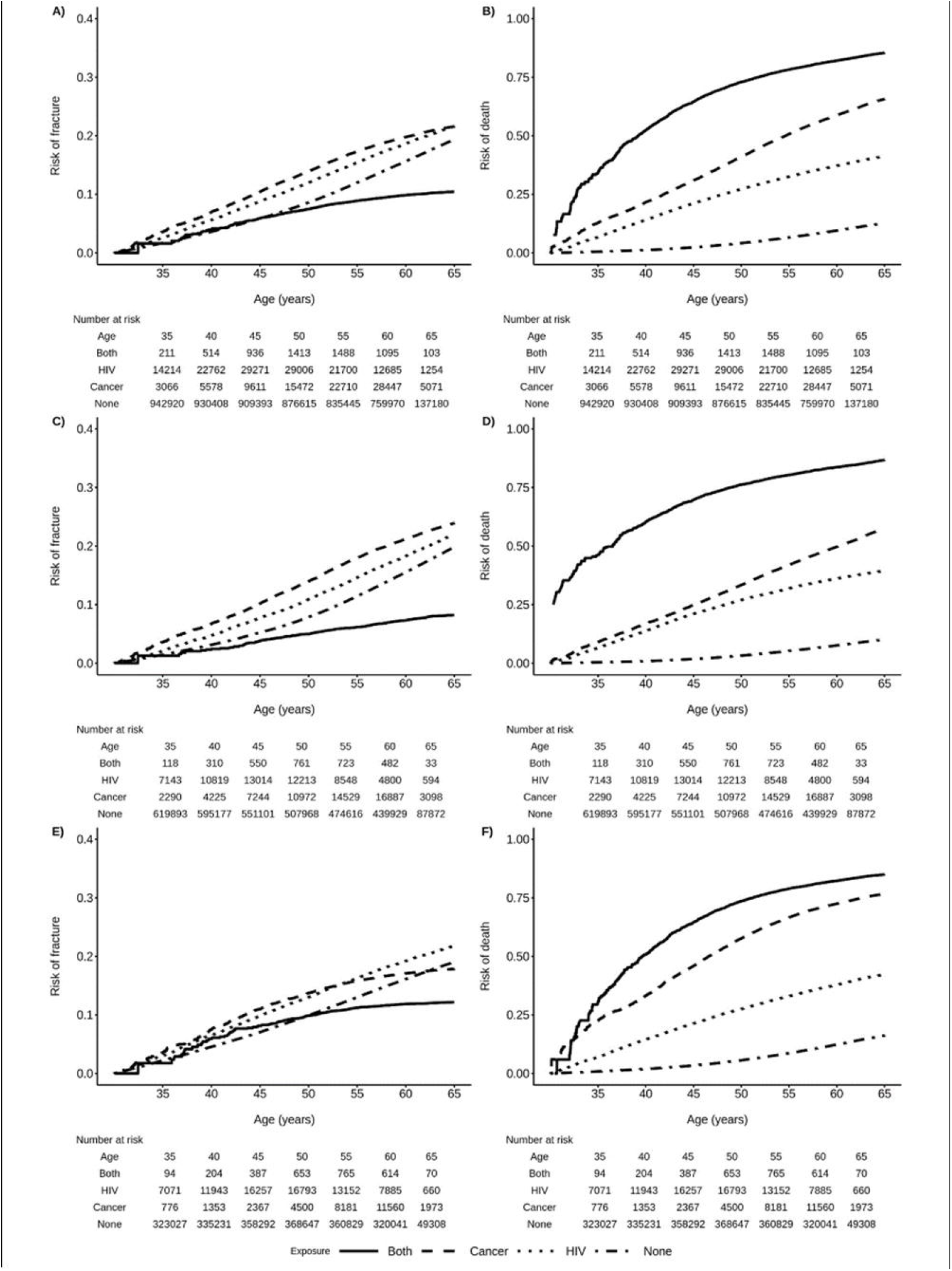
Cumulative incidence of frailty-related fracture and death by sex, age, and HIV and non-AIDS defining cancer (NADC) status among Medicaid Beneficiaries, 2001-2015. A) Risk of first frailty-related fracture; B) Risk of death; C) Risk of first frailty-related fracture among female beneficiaries; D) Risk of death among female beneficiaries; E) Risk of first frailty-related fracture among male beneficiaries; F) Risk of death among male beneficiaries.

Findings were similar when restricting to AIDS-defining cancers, female breast cancer, and lung cancer (Figure S1 A,C,G) and by calendar period (Figure S2). In contrast, cumulative incidence of frailty-related fracture was highest among beneficiaries with both HIV and cancer when restricting to colon cancer and male prostate cancer (Figure S1 E,I). Cumulative incidence of death was substantially lower for male beneficiaries with HIV and prostate cancer (Figure S1 J) than for male beneficiaries with HIV and any NADC (Figure 2F).

## Discussion

Beneficiaries with HIV, NADC, or both conditions had significantly higher incidence rates of frailty-related fracture than beneficiaries without either condition at all ages between 30-64. Beneficiaries with both HIV and NADC had higher incidence of frailty-related fracture than beneficiaries with HIV only. However, the actual risk (i.e., cumulative incidence) of frailty-related fracture was lowest among beneficiaries with HIV and NADC, due to their substantially higher age-specific rates and cumulative incidence of death. This suggests that those with HIV and NADC had a higher rate of fracture when alive but were more likely to die prior to fracture occurring. Findings were generally similar for males and females and by cancer type.

Our finding for increased rates of fracture among beneficiaries with both HIV and NADC was similar to one prior study that reported that PLWH with a recent NADC diagnosis had 1.8 times the rate of fracture compared to PLWH without NADC.^17^ In that study, a recent ADC was not associated with fracture, but was associated with bone disease. In our study, cumulative incidence of frailty-related fracture was lowest among beneficiaries with HIV and ADC and those with HIV and lung cancer, in part because cumulative incidence of death was also highest among these groups. In contrast, cumulative incidence of fracture was substantially higher among male beneficiaries with HIV and prostate cancer and modestly higher among beneficiaries with HIV and colon cancer as compared to other groups. Notably cumulative incidence of death for these cancers was similar or lower than beneficiaries with cancer alone.

Consistent with prior reports, we observed higher incidence of frailty-related fracture among those with HIV or cancer, and among older beneficiaries.^1–5,14,15,25^ Incidence rates of fracture among beneficiaries with HIV were similar to those previously reported in a meta-analysis.^7^ Incidence rates among beneficiaries with cancer were similar to a prior study that evaluated fracture in cancer survivors, approximately 70.6% of whom were under the age of 65, which aligned with the age of our study.^15^

We evaluated the incidence of frailty-related fracture among more than 14 million Medicaid beneficiaries by cancer and HIV status, between 2001 and 2015 across 14 US states, using time-updated measures of HIV and cancer status. Our study reflects the experience of the approximately 40% of PLWH in the US covered by Medicaid. Our large analytic sample enabled us to assess fracture by age, sex, and cumulative incidence for major cancer types, but we had insufficient sample size to report age-specific incidence by cancer type. We lacked cancer-specific information, like the presence of bone metastases,^31^ and HIV-specific information, like CD4 count,^32,33^ which have previously been associated with fracture risk. Our study population included beneficiaries 30-64 years. While fracture risk may be higher in individuals >65 years, only ∼3% of PLWH ≥30 years were >65 during this time period.^34,35^ Thus, our analysis is relevant to the experience of a large proportion of PLWH at risk for frailty-related fracture.

Beneficiaries with HIV and NADC had higher incidence rates of age-specific frailty-related fracture than beneficiaries with HIV alone, including at younger ages. Incidence of death was also exceptionally high among this group, which decreased the observed risk of frailty-related fracture. More work is needed to assure that PLWH have access high quality HIV care and care across the cancer continuum (e.g. primary prevention through treatment) to reduce cancer burden and improve overall survival. Future work should characterize fracture risk by cancer type to better inform interventions for fracture prevention among PLWH with cancer. Furthermore, clinical providers should be aware of high incidence rates of fracture among PLWH with cancer including at young ages.

## Data Availability

The data that support the findings of this study are available from Research Data Assistance Center (ResDAC). Restrictions apply to the availability of these data, which were used under license for this study.

https://resdac.org/

## Funding

This work was supported by NIH grants R01CA250851, R01AI170240, U01AI069918, and P30CA006973. CEJ was also supported by a grant from the American Cancer Society (Grant No. RSG-18-147-01). This research was also funded in part by a 2018 and 2021 developmental grant from the Johns Hopkins University Center for AIDS Research, an NIH funded program (P30AI094189), which is supported by the following NIH Co-Funding and Participating Institutes and Centers: NIAID, NCI, NICHD, NHLBI, NIDA, NIA, NIGMS, NIDDK, NIMHD.

### Role of funding source

The content is solely the responsibility of the authors and does not necessarily represent the official views of the NIH.

## Notes

### Competing Interest Statement

Funding for the study was provided in part by the American Cancer Society. Corinne Joshu is a paid grant reviewer for the American Cancer Society. This arrangement has been reviewed and approved by the Johns Hopkins University in accordance with its conflict-of-interest policies.

### Funding Statement

This work was funded in part by NIH grants R01CA250851, R01AI170240, U01AI069918, and P30CA006973. CEJ was also supported by a grant from the American Cancer Society (Grant No. RSG-18-147-01). This research was also funded in part by a 2018 and 2021 developmental grant from the Johns Hopkins University Center for AIDS Research, an NIH funded program (P30AI094189), which is supported by the following NIH Co-Funding and Participating Institutes and Centers: NIAID, NCI, NICHD, NHLBI, NIDA, NIA, NIGMS, NIDDK, NIMHD. The content is solely the responsibility of the authors and does not necessarily represent the official views of the NIH.

### Author Declarations

The Johns Hopkins Bloomberg School of Public Health Institutional Review Board approved and waived the requirement for informed consent for this analysis of a Limited Data Set, which contains protected health information including dates of birth, death, and healthcare services.

