## Supplemental Tables and Figures for "Incidence of frailty-related fracture among Medicaid beneficiaries living with HIV and cancer: A cohort study"

Table S1. The International Classification of Diseases 9th edition (ICD-9) codes used to identify HIV diagnoses, cancer diagnoses and fracture diagnosis.

| Variable | | ICD-9[a] |
|---|---|---|
| Human Immunodeficiency Virus | | 042-044, 079.53, 795.71, V08 |
| AIDS-defining cancers | | |
| | Cervical | 180.X |
| | Kaposi's sarcoma | 176.X |
| | Non-Hodgkin's lymphoma | 200.X, 202.X |
| Non-AIDS-defining cancers | | |
| | Anal[b] | 154.2, 154.3 |
| | Bladder | 188.X |
| | Brain | 191.X |
| | Breast | 174.X, 175.X |
| | Colon | 153.X |
| | Esophagus | 150.X |
| | Head & neck | 140.X-149.X, 160.X, 161.X |
| | Hodgkin's lymphoma[b] | 201.X |
| | Kidney | 189.X |
| | Larynx | 161.X |
| | Leukemia | 204.X-208.X |
| | Liver[b] | 155.X |
| | Lung | 162.2-162.5, 162.8, 162.9 |
| | Melanoma | 172.X |
| | Myeloma | 203.X |
| | Oropharynx[b] | 146.X |
| | Ovary | 183.X |
| | Pancreas | 157.X |
| | Penile[b] | 187.X |
| | Prostate | 185.X |
| | Rectal | 154.0, 154.1 |
| | Stomach[b] | 151.X |
| | Uterine | 179.X, 182.X |
| | Vaginal/vulvar[b] | 184.X |
| Frailty-related fractures | | |
| | Hip fracture | 733.14, 820.X |
| | Vertebral | 805.X |
| | Pelvic | 808.X |
| | Wrist | 813.X |

Note: [a]A code ending with ".X" indicates a wildcard. Any number could appear after the decimal place. [b]Infection-related non-AIDS-defining cancer. In addition to oropharyngeal cancers, head-neck cancers with a code 141.0, 141.6, and 149.1 were classified as infection-related.

Table S2. Baseline characteristics of Medicaid beneficiaries by HIV status, 2001-2015

| Characteristics | No HIV | HIV |
|---|---|---|
| N | 14,554,711 | 159,188 |
| Age (year) (Median (Q25, Q75)) | 43.5 (35.2, 53.8) | 43.9 (37.4, 51.0) |
| Female, N (%) | 8,314,883 (57.1%) | 55,316 (34.7%) |
| Race/ethnicity, N (%) | | |
| non-Hispanic white | 6,188,473 (42.5%) | 39,447 (24.8%) |
| non-Hispanic Black | 2,999,662 (20.6%) | 76,046 (47.8%) |
| Hispanic | 2230468 (15.3%) | 14892 (9.4%) |
| Other/Missing | 3136108 (21.5%) | 28803 (18.1%) |
| State, N (%) | | |
| Alabama | 186,581 (1.3%) | 1,694 (1.1%) |
| California | 4,118,455 (28.3%) | 26,724 (16.8%) |
| Colorado | 206,996 (1.4%) | 713 (0.4%) |
| Florida | 922,304 (6.3%) | 18,551 (11.7%) |
| Georgia | 445,851 (3.1%) | 6,799 (4.3%) |
| Illinois | 1,196,744 (8.2%) | 8,595 (5.4%) |
| Massachusetts | 748,713 (5.1%) | 6,858 (4.3%) |
| Maryland | 389,126 (2.7%) | 6,631 (4.2%) |
| North Carolina | 451,637 (3.1%) | 6,228 (3.9%) |
| New York | 2,482,128 (17.1%) | 56,134 (35.3%) |
| Ohio | 930,941 (6.4%) | 4,321 (2.7%) |
| Pennsylvania | 860,089 (5.9%) | 4,391 (2.8%) |
| Texas | 842,897 (5.8%) | 7,971 (5.0%) |
| Washington | 772,249 (5.3%) | 3,578 (2.2%) |

Note: HIV, human immunodeficiency virus; SD, standard deviation; N, number; [a]Other included American Indian or Alaska native, Asian, Native Hawaiian or other Pacific Islander, more than one race, without indication of Hispanic ethnicity.

Table S3. Crude incidence rates of first frailty-related fracture and death by HIV and Non-AIDS defining cancer (NADC) status and age

| | Both HIV and NADC | | Only HIV | | Only NADC | | No HIV and NADC | |
|---|---|---|---|---|---|---|---|---|
| Age Group | Events/PYs | IR (95% CI) | Events/PYs | IR (95% CI) | Events/PYs | IR (95% CI) | Events/PYs | IR (95% CI) |
| 30-44 | | | | | | | | |
| Fracture | 60/ 4,836 | 1.24 (0.96, 1.59) | 2,048/ 276,583 | 0.74 (0.71, 0.77) | 670/ 67,705 | 0.99 (0.92, 1.07) | 57,125/ 1,4076,918 | 0.41 (0.40, 0.41) |
| Death | 399/ 4,836 | 8.25 (7.47, 9.09) | 4,810/ 276,583 | 1.74 (1.69, 1.79) | 1,802/ 67,705 | 2.66 (2.54, 2.79) | 23,035/ 14,076,918 | 0.16 (0.16, 0.17) |
| 45-49 | | | | | | | | |
| Fracture | 77/ 4,717 | 1.63 (1.30, 2.03) | 1,468/ 149,156 | 0.98 (0.93, 1.04) | 842/ 62,027 | 1.36 (1.27, 1.45) | 27,158/ 4,448,093 | 0.61 (0.60, 0.62) |
| Death | 409/ 4,717 | 8.67 (7.86, 9.54) | 2,774/ 149,156 | 1.86 (1.79, 1.93) | 2,455/ 62,027 | 3.96 (3.80, 4.12) | 17,561/ 4,448,093 | 0.39 (0.39, 0.40) |
| 50-54 | | | | | | | | |
| Fracture | 134/ 5,905 | 2.27 (1.91, 2.68) | 1,580/ 128,949 | 1.23 (1.17, 1.29) | 1,696/ 96,220 | 1.76 (1.68, 1.85) | 34,517/ 4,307,206 | 0.80 (0.79, 0.81) |
| Death | 508/ 5,905 | 8.60 (7.88, 9.38) | 2,433/ 128,949 | 1.89 (1.81, 1.96) | 4,932/ 96,220 | 5.13 (4.98, 5.27) | 25,084/ 4,307,206 | 0.58 (0.58, 0.59) |
| 55-59 | | | | | | | | |
| Fracture | 128/ 5,434 | 2.36 (1.97, 2.79) | 1,159/ 86,418 | 1.34 (1.27, 1.42) | 2,442/ 129,940 | 1.88 (1.81, 1.95) | 37,730/ 4,026,119 | 0.94 (0.93, 0.95) |
| Death | 502/ 5,434 | 9.24 (8.46, 10.07) | 1,671/ 86,418 | 1.93 (1.84, 2.03) | 7,952/ 129,940 | 6.12 (5.99, 6.26) | 30,421/ 4,026,119 | 0.76 (0.75, 0.76) |
| 60-64 | | | | | | | | |
| Fracture | 65/ 3,087 | 2.11 (1.64, 2.67) | 728/ 49,362 | 1.47 (1.37, 1.58) | 2,712/ 125,577 | 2.16 (2.08, 2.24) | 37,759/ 3,609,120 | 1.05 (1.04, 1.06) |
| Death | 380/ 3,087 | 12.31 (11.12, 13.60) | 1,001/ 49,362 | 2.03 (1.91, 2.16) | 10,303/ 125,577 | 8.20 (8.05, 8.36) | 31,546/ 3,609,120 | 0.87 (0.86, 0.88) |

IR, incidence rate per 100 person-years; PY, person-year.

Table S4. Crude incidence rates of first frailty-related fracture and death by HIV and Non-AIDS defining cancer (NADC) status, age, and sex

| | Both HIV and NADC | | Only HIV | | Only NADC | | No HIV and NADC | |
|---|---|---|---|---|---|---|---|---|
| Age Group | Events/PYs | IR (95% CI) | Events/PYs | IR (95% CI) | Events/PYs | IR (95% CI) | Events/PYs | IR (95% CI) |
| *Female* | | | | | | | | |
| 30-44 | | | | | | | | |
| Fracture | 29/2,663 | 1.09 (0.74, 1.54) | 858/133,202 | 0.64 (0.60, 0.69) | 462/51,277 | 0.90 (0.82, 0.99) | 32,427/9,108,129 | 0.36 (0.35, 0.36) |
| Death | 210/2,663 | 7.89 (6.87, 9.01) | 2,285/133,202 | 1.72 (1.65, 1.79) | 1,087/51,277 | 2.12 (2.00, 2.25) | 10,959/9,108,129 | 0.12 (0.12, 0.12) |
| 45-49 | | | | | | | | |
| Fracture | 33/2,441 | 1.35 (0.95, 1.88) | 631/64,809 | 0.97 (0.90, 1.05) | 585/45,385 | 1.29 (1.19, 1.40) | 15,255/2,632,110 | 0.58 (0.57, 0.59) |
| Death | 191/2,441 | 7.82 (6.77, 8.99) | 1,148/64,809 | 1.77 (1.67, 1.88) | 1,339/45,385 | 2.95 (2.80, 3.11) | 8,335/2,632,110 | 0.32 (0.31, 0.32) |
| 50-54 | | | | | | | | |
| Fracture | 59/2,855 | 2.07 (1.59, 2.65) | 671/52,425 | 1.28 (1.19, 1.38) | 1,115/64,482 | 1.73 (1.63, 1.83) | 20,417/2,460,542 | 0.83 (0.82, 0.84) |
| Death | 209/2,855 | 7.32 (6.38, 8.37) | 900/52,425 | 1.72 (1.61, 1.83) | 2,421/64,482 | 3.75 (3.61, 3.91) | 11,271/2,460,542 | 0.46 (0.45, 0.47) |
| 55-59 | | | | | | | | |
| Fracture | 66/2,399 | 2.75 (2.14, 3.48) | 491/33,344 | 1.47 (1.35, 1.61) | 1,516/79,738 | 1.90 (1.81, 2.00) | 23,473/2,297,761 | 1.02 (1.01, 1.03) |
| Death | 191/2,399 | 7.96 (6.89, 9.15) | 577/33,344 | 1.73 (1.59, 1.88) | 3,606/79,738 | 4.52 (4.38, 4.67) | 13,620/2,297,761 | 0.59 (0.58, 0.60) |
| 60-64 | | | | | | | | |
| Fracture | 37/1,178 | 3.14 (2.24, 4.28) | 334/19,386 | 1.72 (1.55, 1.92) | 1,650/72,044 | 2.29 (2.18, 2.40) | 25,192/2,146,867 | 1.17 (1.16, 1.19) |
| Death | 114/1,178 | 9.68 (8.02, 11.58) | 306/19,386 | 1.58 (1.41, 1.76) | 4,609/72,044 | 6.40 (6.21, 6.58) | 14,776/2,146,867 | 0.69 (0.68, 0.70) |
| *Male* | | | | | | | | |
| 30-44 | | | | | | | | |
| Fracture | 31/2,172 | 1.43 (0.99, 2.00) | 1,190/143,380 | 0.83 (0.78, 0.88) | 208/16,427 | 1.27 (1.10, 1.45) | 24,698/4,968,788 | 0.50 (0.49, 0.50) |
| Death | 189/2,172 | 8.70 (7.52, 10.01) | 2,525/143,380 | 1.76 (1.69, 1.83) | 715/16,427 | 4.35 (4.04, 4.68) | 12,076/4,968,788 | 0.24 (0.24, 0.25) |
| 45-49 | | | | | | | | |
| Fracture | 44/2,276 | 1.93 (1.42, 2.57) | 837/84,347 | 0.99 (0.93, 1.06) | 257/16,642 | 1.54 (1.36, 1.74) | 11,903/1,815,983 | 0.66 (0.64, 0.67) |
| Death | 218/2,276 | 9.58 | 1,626/84,347 | 1.93 | 1,116/16,642 | 6.71 | 9,226/1,815,983 | 0.51 |

| | | | | | | | | |
|---|---|---|---|---|---|---|---|---|
| | | (8.37, 10.91) | | (1.84, 2.02) | | (6.32, 7.11) | | (0.50, 0.52) |
| 50-54 | | | | | | | | |
| Fracture | 75/3,050 | 2.46 (1.95, 3.06) | 909/76,524 | 1.19 (1.11, 1.27) | 581/31,738 | 1.83 (1.69, 1.98) | 14,100/1,846,664 | 0.76 (0.75, 0.78) |
| Death | 299/3,050 | 9.80 (8.74, 10.96) | 1,533/76,524 | 2.00 (1.90, 2.11) | 2,511/31,738 | 7.91 (7.61, 8.23) | 13,813/1,846,664 | 0.75 (0.74, 0.76) |
| 55-59 | | | | | | | | |
| Fracture | 62/3,035 | 2.04 (1.58, 2.60) | 668/53,074 | 1.26 (1.17, 1.36) | 926/50,202 | 1.84 (1.73, 1.97) | 14,257/1,728,358 | 0.82 (0.81, 0.84) |
| Death | 311/3,035 | 10.25 (9.16, 11.44) | 1,094/53,074 | 2.06 (1.94, 2.19) | 4,346/50,202 | 8.66 (8.40, 8.92) | 16,801/1,728,358 | 0.97 (0.96, 0.99) |
| 60-64 | | | | | | | | |
| Fracture | 28/1,908 | 1.47 (0.99, 2.09) | 394/29,976 | 1.31 (1.19, 1.45) | 1,062/53,533 | 1.98 (1.87, 2.11) | 12,567/1,462,253 | 0.86 (0.84, 0.87) |
| Death | 266/1,908 | 13.94 (12.34, 15.69) | 695/29,976 | 2.32 (2.15, 2.50) | 5,694/53,533 | 10.64 (10.36, 10.92) | 16,770/1,462,253 | 1.15 (1.13, 1.16) |

IR, incidence rate per 100 person-years; PY, person-year.

Table S5. Proportion (%) of frailty-related fractures by age, body location, sex, and HIV and Non-AIDS defining cancer (NADC) status

| Age Group | | Proportion | | | |
|---|---|---|---|---|---|
| | | Both HIV and NADC | Only HIV | Only NADC | No HIV and NADC |
| *Female* | | | | | |
| 30-44 | | | | | |
| | Hip | 48.28 | 17.81 | 27.29 | 11.23 |
| | Pelvic | 13.79 | 8.96 | 9.59 | 7.33 |
| | Vertebral | 10.34 | 24.97 | 32.41 | 31.29 |
| | Wrist | 27.59 | 48.26 | 30.70 | 50.14 |
| 45-49 | | | | | |
| | Hip | 36.36 | 20.92 | 23.62 | 14.95 |
| | Pelvic | 9.09 | 9.23 | 9.55 | 7.06 |
| | Vertebral | 21.21 | 23.69 | 35.85 | 29.39 |
| | Wrist | 33.33 | 46.15 | 30.99 | 48.59 |
| 50-54 | | | | | |
| | Hip | 25.40 | 22.77 | 25.68 | 17.09 |
| | Pelvic | 9.52 | 8.76 | 7.77 | 6.76 |
| | Vertebral | 17.46 | 22.92 | 32.14 | 27.68 |
| | Wrist | 47.62 | 45.55 | 34.41 | 48.46 |
| 55-59 | | | | | |
| | Hip | 36.23 | 24.40 | 27.54 | 19.92 |
| | Pelvic | 4.35 | 9.72 | 8.08 | 6.95 |
| | Vertebral | 27.54 | 24.80 | 34.26 | 27.26 |
| | Wrist | 31.88 | 41.07 | 30.12 | 45.86 |
| 60-64 | | | | | |
| | Hip | 28.95 | 27.38 | 32.13 | 22.97 |
| | Pelvic | 15.79 | 9.80 | 8.21 | 7.16 |
| | Vertebral | 21.05 | 26.51 | 35.62 | 27.92 |
| | Wrist | 34.21 | 36.31 | 24.04 | 41.95 |
| *Male* | | | | | |
| 30-44 | | | | | |
| | Hip | 29.03 | 20.20 | 28.44 | 14.53 |
| | Pelvic | 0 | 9.04 | 6.16 | 7.52 |
| | Vertebral | 38.71 | 24.35 | 35.55 | 32.59 |
| | Wrist | 32.26 | 46.42 | 29.86 | 45.36 |
| 45-49 | | | | | |
| | Hip | 31.82 | 24.36 | 27.38 | 17.84 |
| | Pelvic | 9.09 | 8.08 | 5.70 | 7.39 |
| | Vertebral | 29.55 | 25.17 | 40.30 | 33.92 |
| | Wrist | 29.55 | 42.38 | 26.62 | 40.85 |
| 50-54 | | | | | |
| | Hip | 35.06 | 25.16 | 31.50 | 21.91 |
| | Pelvic | 3.90 | 7.71 | 6.67 | 7.52 |
| | Vertebral | 37.66 | 25.05 | 37.17 | 34.59 |
| | Wrist | 23.38 | 42.08 | 24.67 | 35.98 |
| 55-59 | | | | | |
| | Hip | 41.27 | 32.40 | 32.32 | 25.70 |
| | Pelvic | 6.35 | 6.92 | 6.53 | 7.38 |
| | Vertebral | 23.81 | 24.74 | 41.16 | 34.93 |
| | Wrist | 28.57 | 35.94 | 20.00 | 31.99 |
| 60-64 | | | | | |
| | Hip | 35.71 | 39.70 | 36.44 | 30.73 |
| | Pelvic | 3.57 | 5.21 | 8.31 | 7.50 |
| | Vertebral | 39.29 | 26.30 | 38.45 | 33.50 |
| | Wrist | 21.43 | 28.78 | 16.80 | 28.26 |

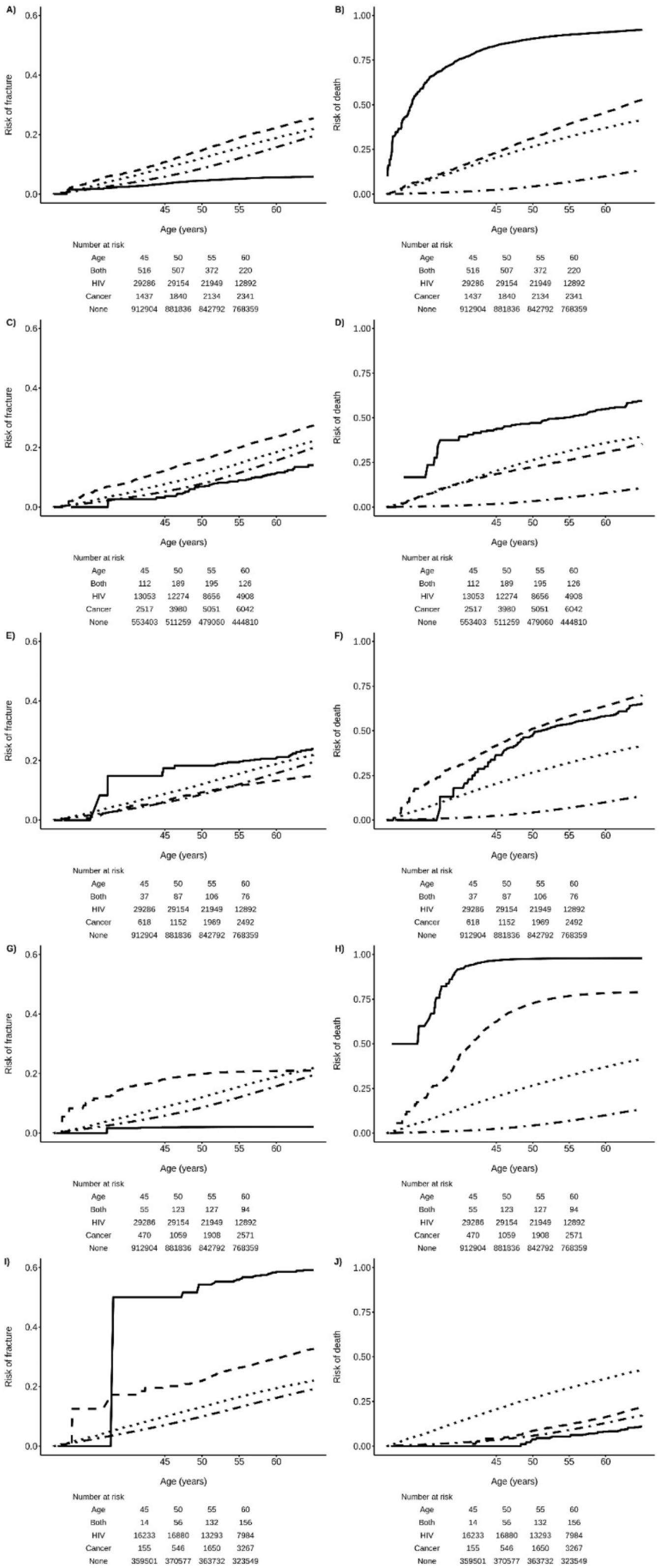


Figure S1. Cumulative incidence of frailty-related fracture and death by sex, age, and HIV and cancer type among Medicaid Beneficiaries, 2001-2015. A) Risk of first frailty-related fracture among beneficiaries with AIDS-defining cancer; B) Risk of death among beneficiaries with AIDS-defining cancer; C) Risk of first frailty-related fracture among female beneficiaries with breast cancer; D) Risk of death among female beneficiaries with breast cancer; E) Risk of first frailty-related fracture among beneficiaries with colon cancer; F) Risk of death among beneficiaries with colon cancer; G) Risk of first frailty-related fracture among beneficiaries with lung cancer; H) Risk of death among beneficiaries with lung cancer; I) Risk of first frailty-related fracture among male beneficiaries with prostate cancer; J) Risk of death among male beneficiaries with prostate cancer.

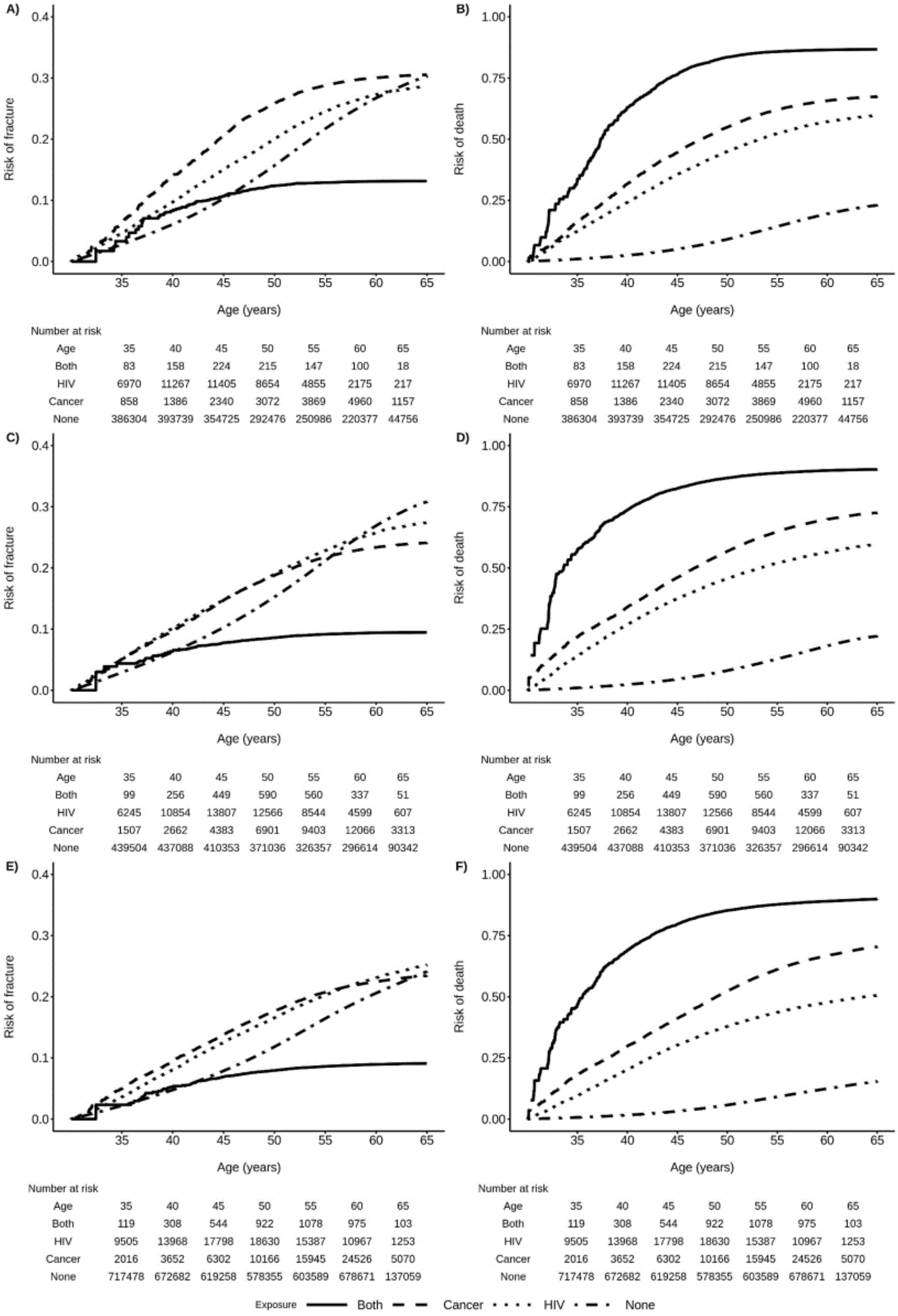


Number at risk (A)

| Age | 35 | 40 | 45 | 50 | 55 | 60 | 65 |
|---|---|---|---|---|---|---|---|
| Both | 83 | 158 | 224 | 215 | 147 | 100 | 18 |
| HIV | 6970 | 11267 | 11405 | 8654 | 4855 | 2175 | 217 |
| Cancer | 858 | 1386 | 2340 | 3072 | 3869 | 4960 | 1157 |
| None | 386304 | 393739 | 354725 | 292476 | 250986 | 220377 | 44756 |

Number at risk (B)

| Age | 35 | 40 | 45 | 50 | 55 | 60 | 65 |
|---|---|---|---|---|---|---|---|
| Both | 83 | 158 | 224 | 215 | 147 | 100 | 18 |
| HIV | 6970 | 11267 | 11405 | 8654 | 4855 | 2175 | 217 |
| Cancer | 858 | 1386 | 2340 | 3072 | 3869 | 4960 | 1157 |
| None | 386304 | 393739 | 354725 | 292476 | 250986 | 220377 | 44756 |

Number at risk (C)

| Age | 35 | 40 | 45 | 50 | 55 | 60 | 65 |
|---|---|---|---|---|---|---|---|
| Both | 99 | 256 | 449 | 590 | 560 | 337 | 51 |
| HIV | 6245 | 10854 | 13807 | 12566 | 8544 | 4599 | 607 |
| Cancer | 1507 | 2662 | 4383 | 6901 | 9403 | 12066 | 3313 |
| None | 439504 | 437088 | 410353 | 371036 | 326357 | 296614 | 90342 |

Number at risk (D)

| Age | 35 | 40 | 45 | 50 | 55 | 60 | 65 |
|---|---|---|---|---|---|---|---|
| Both | 99 | 256 | 449 | 590 | 560 | 337 | 51 |
| HIV | 6245 | 10854 | 13807 | 12566 | 8544 | 4599 | 607 |
| Cancer | 1507 | 2662 | 4383 | 6901 | 9403 | 12066 | 3313 |
| None | 439504 | 437088 | 410353 | 371036 | 326357 | 296614 | 90342 |

Number at risk (E)

| Age | 35 | 40 | 45 | 50 | 55 | 60 | 65 |
|---|---|---|---|---|---|---|---|
| Both | 119 | 308 | 544 | 922 | 1078 | 975 | 103 |
| HIV | 9505 | 13968 | 17798 | 18630 | 15387 | 10967 | 1253 |
| Cancer | 2016 | 3652 | 6302 | 10166 | 15945 | 24526 | 5070 |
| None | 717478 | 672682 | 619258 | 578355 | 603589 | 678671 | 137059 |

Number at risk (F)

| Age | 35 | 40 | 45 | 50 | 55 | 60 | 65 |
|---|---|---|---|---|---|---|---|
| Both | 119 | 308 | 544 | 922 | 1078 | 975 | 103 |
| HIV | 9505 | 13968 | 17798 | 18630 | 15387 | 10967 | 1253 |
| Cancer | 2016 | 3652 | 6302 | 10166 | 15945 | 24526 | 5070 |
| None | 717478 | 672682 | 619258 | 578355 | 603589 | 678671 | 137059 |

Figure S2. Cumulative incidence of frailty-related fracture and death by sex, age, HIV and Non-AIDS defining cancer, and calendar period among Medicaid Beneficiaries, 2001-2015. A) Risk of first frailty-related fracture among beneficiaries in 2001-2005; B) Risk of death among beneficiaries in 2001-2005; C) Risk of first frailty-related fracture among female beneficiaries in 2006-2010; D) Risk of death among female beneficiaries in 2006-2010; E) Risk of first frailty-related fracture among beneficiaries in 2011-2015; F) Risk of death among beneficiaries in 2011-2015.
